# Projecting trends in the disease burden of adult edentulism in China between 2020 and 2030: a systematic study based on the global burden of disease

**DOI:** 10.1101/2023.12.22.23300444

**Authors:** Xiaofeng Qin, Li Chen, Xihua Yuan, Dan Lin, Qiulin Liu, Xiaojuan Zeng, Fei Ma

**Affiliations:** Guangxi Key Laboratory of Oral and Maxillofacial Rehabilitation and Reconstruction, College of Stomatology, Hospital of Stomatology, Guangxi Medical University, Nanning, China; Department of Dermatology and Venerology, The Second Affiliated Hospital, Guangxi Medical University, Nanning, China

**Keywords:** Edentulism, GBD, incidence, prevalence, YLDs

## Abstract

**PURPOSE:** This study was based on the Global Burden of Disease (GBD) database and aimed to analyze the trend of disease burden for edentulism in Chinese adults between 1990 and 2030, and to provide valuable information for the development of more effective management and preventive measures.

**METHODS:** Data on Chinese adults with edentulism from 1990 to 2019 was analyzed using GHDx data. Descriptive analyses were used to analyze changes in the prevalence and burden of edentulism, gender and age distribution between 1990 and 2019. In addition, we used an autoregressive integrated moving average (ARIMA) model to predict the trend of disease burden for Chinese adults with edentulism between 2020 and 2030.

**RESULTS:** The incidence, prevalence, and rate of YLDs in adults with edentulism in China showed an increasing trend from 1990 to 2019. In 2019, the incidence was 251.20 per 100,000, the prevalence was 4512.78 per 100,000, and the YLDs were 123.44 per 100,000, marking increases of 20.58%, 94.18%, and 93.12% from 1990. Males experienced a higher increase than females. However, the standardized rates decreased over the same period. The ARIMA model predicts a subsequent upward and then downward trend for all indicators between 2019 and 2030, except for the standardized incidence rate which remained essentially unchanged. Specifically, the incidence is predicted to decrease from 388.93 to 314.40 per 100,000, prevalence from 4512.78 to 3049.70 per 100,000, and YLDs from 123.44 to 103.44 per 100,000. The standardized prevalence and YLDs rates are also expected to decrease.

**CONCLUSION:** The burden of edentulism in China is projected to show an increasing trend from 2020 to 2022 and a decreasing trend from 2023 to 2030. Despite the decline in the burden of disease associated with edentulism in China, many problems remain to be solved.

## 1. Introduction

The case definition of edentulism includes any individual with zero remaining permanent teeth; toothlessness of infancy is not included.. Missing teeth often generate multiple adverse effects; on the one hand, this will lead to reduced chewing ability and limited food choices, especially for the elderly. Patients with missing teeth tend to avoid choosing fruits, vegetables, nuts and other tougher but healthy food that is needed by the body, thus resulting in an imbalance in nutritional intake [1–5]. On the other hand, missing teeth will also lead to alveolar bone resorption, shortening of the vertical distance of the lower third of the face, relaxation of the facial muscles, lip and cheek subsidence, and other changes in appearance, thus resulting in an aging face, unclear pronunciation, along with psychological and social disorders [6]. In addition, missing teeth can affect systemic diseases and may increase the risk of stroke, myocardial ischemia, coronary heart disease, cognitive impairment and other diseases [7–10].

Missing teeth place a heavy economic burden on society. The economic burden of oral disease is the fourth highest of all diseases in most industrialized countries around the world; in 2010, the global economic burden of oral disease was $442 billion, of which the direct cost of treatment was $298 billion, or approximately 4.6% of global health expenditure [11]. Between 2010 and 2015, indirect costs increased by 21.0% as a result of oral diseases, with 67.0% of lost productivity attributable to severe tooth loss (fewer than nine permanent teeth) [12]. As the most populous country in the world, research predicts that the proportion of the elderly population (aged 65 years and above) in China will reach 19.25% by 2030 [13], and that this country will enter a stage of heavy ageing, where the burden of diseases caused by missing teeth in the elderly will increase and become a social problem that cannot be ignored.

In a previous study, we found that the incidence, prevalence, and years lived with disability (YLDs) of Chinese adults with missing teeth were followed an increasing trend between 1990 and 2019, while the standardized incidence, prevalence, and YLDs showed a decreasing trend [14]; this opposing trend was due to the increasing aging of the population. As population aging in China becomes further aggravated, it is important to investigate whether future changes in the trends of the rate and the standardized rate are consistent. Therefore, in this study, we constructed an autoregressive integrated moving average (ARIMA) model to predict the disease burden in China between 2020 and 2030. This can not only provide a scientific basis for the scientific formulation of national public health policies and the rational allocation of medical and health resources, but also provide a reference for health administrations to determine the priority areas of disease prevention and control, and the formulation of strategies for the prevention and control of chronic diseases.

## 2 Materials and Methods

### 2.1 Data sources

The incidence, prevalence and YLDs associated with missing teeth and their corresponding age-standardized rates (ASRs) in China between 1990 and 2019 were obtained from the Global Health Data Exchange query tool (http://ghdx.healthdata.org/gbd-results-tool). The disability weighting for missing teeth in the GBD 2019 study was 0.067 (0.045–0.095) and can be used to estimate the Disability-adjusted life years (DALYs) for missing teeth. Age-standardized rates for missing teeth were based on the GBD 2019 global age-standardized population. DALYs are a crucial demographic indicator in the GBD and represent the sum of YLLs and YLDs. Deaths caused directly by oral disease are uncommon. Therefore, only YLDs were used in this study. YLDs are calculated by summing the frequency (prevalence), severity (weight of disability), and duration of a condition. The reliability of GBD data has been confirmed by previous studies [14–21].

### 2.2. Definition of edentulism

The case definition of edentulism includes any individual with no remaining permanent teeth, excluding edentulousness in infancy. This disease is evaluated by quantifying its incidence and estimating the primary sequelae; specifically, asymptomatic and symptomatic edentulism resulting in significant difficulty in eating meat, fruits, and vegetables. A small body of evidence suggests that edentulism predisposes individuals to an increased risk of ischemic cardiovascular events, including myocardial infarction and stroke. These sparse data have been incorporated into models that estimate the excess mortality in individuals with complete tooth loss. However, as this association is considered ecological rather than causal, tooth loss was not estimated as an underlying cause of death. Therefore, this was included in the analysis of risk factors for cardiovascular disease. The International Classification of Diseases (ICD) codes for tooth loss mapped to the Global Burden of Disease list of causes are K08.0-K08.499 for ICD10; and 525.0-525.19, 525.4-525.54 for ICD9[18].

### 2.3 Statistical analyses

An ARIMA model is a type of differential, integral, moving average and autoregressive model, also known as an ‘integral moving average autoregressive model’, and represents a model that is commonly used to use time series of data for forecasting analysis. In the ARIMA model (*p, d, q*), AR is ‘autoregressive’, *p* is the number of autoregressive terms, MA is ‘moving average’, *q* is the number of terms in the moving average, and d is the number of differences (orders) that provide a smooth series [22]. The optimal model was automatically filtered by Akaike information criterion (AIC) and Bayesian information criterion (BIC)[23]. In this study, the ARIMA model was used to analyze the burden of disease based on the trend for disease burden and to predict the burden of disease associated with edentulism in China between 2020 and 2030. All analyses and data visualization were implemented using R3.6.0 software. The accuracy of the model was determined by mean absolute error (MAE), root mean square error (RMSE), mean square error (MSE), mean absolute percentage error (MAPE), symmetric mean absolute percentage error (SMAPE) and other indicators for validation; a smaller MAPE value indicated a smaller error.

## 3. Results

### 3.1 Distribution of incidence, prevalence, and YLDs rate of missing teeth by sex and age in China (2019)

The age and sex distribution of the burden of disease for edentulism in 2019 is shown in Table 1. The incidence, prevalence and YLD rates for edentulism were lowest in the 15–49 year group; there was a rapid upwards trend after the age of 50 years, and the highest rates were evident in the oldest age group (70 years and above; an incidence of 1,908.06/100,000, a prevalence of 28,311.20/100,000, and a YLD rate of 751.03/100,000). Comparison of the disease burdens for edentulism between different genders revealed that the incidence, prevalence and YLD rates were higher in females than in males.

**Table 1.** The age and sex distribution of the burden of disease for edentulism in China, 2019.

| Age | Incidence |  |  | Prevalence |  |  | YLDs |  |  |
| --- | --- | --- | --- | --- | --- | --- | --- | --- | --- |
|  | Both | Male | Female | Both | Male | Female | Both | Male | female |
| All ages | 388.93 | 301.64 | 479.63 | 4512.78 | 3205.16 | 5871.56 | 123.44 | 88.52 | 159.72 |
| 15-49 years | 78.59 | 57.59 | 100.71 | 884.38 | 586.43 | 1198.07 | 25.53 | 17.02 | 34.49 |
| 50-69 years | 787.63 | 612.69 | 962.66 | 7386.47 | 5559.47 | 9214.56 | 206.26 | 156.32 | 256.23 |
| 70+ years | 1908.06 | 1712.15 | 2072.33 | 28311.20 | 21947.90 | 33646.73 | 751.03 | 589.57 | 886.41 |

YLDs: years lived with disability

### 3.2 Forecasts for the incidence, prevalence, and YLDs rates of edentulism in China (2020– 2030)

Predictions of the incidence, prevalence, and YLDs of edentulism, and the incidence, prevalence, and YLDs of standardized edentulism, in Chinese adults with missing teeth from 2020 to 2030 are shown in Table 2 and Figure 1. Between 2020 and 2030, the incidence and standardized rates of edentulism in Chinese adults with missing teeth show a tendency to first increase and then decrease. Projections showed that between 2019 and 2030, the prevalence of missing teeth in China will decrease from 388.93/100,000 to 314.40 /100,000, a reduction of 19.16% when compared with 2019. The standardized prevalence of missing teeth in China is projected to increase slightly from 272.15/100,000 to 277.48/100,000, an increase of 1.79% when compared with 2019. The prevalence of missing teeth in China is projected to decrease from 4512.78/100,000 to 3049.70/100,000, a reduction of 32.42% when compared with 2019. The standardized prevalence of missing teeth in China is projected to decrease from 3327.48/100,000 to 2043.77/100,000, a reduction of 38.58% when compared with 2019. The YLDs of missing teeth in China is projected to decrease from 123.44/100,000 to 103.44/100,000, a reduction of 16.20% compared with 2019.

**Figure 1.**
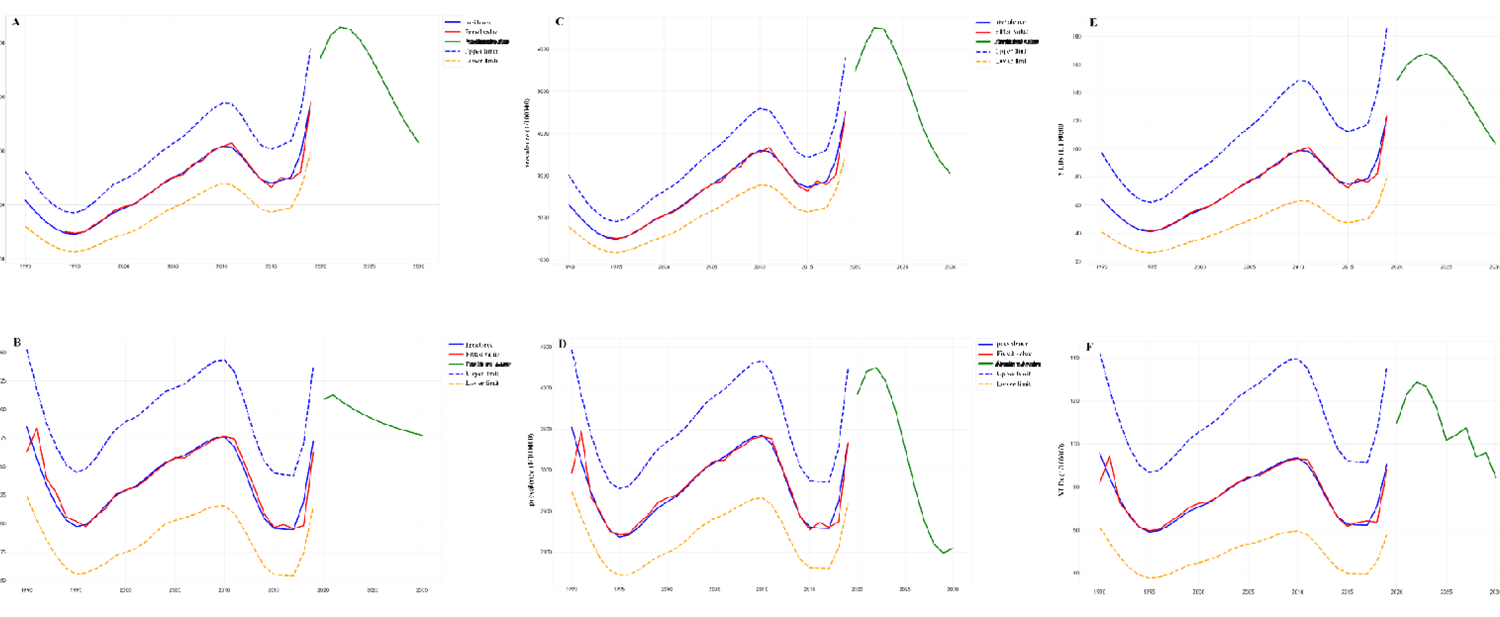
Projected disease burden for edentulism in China between 2020 and 2030. (A) Incidence. (B) Standardized incidence. (C) Prevalence. (D) Standardized prevalence. (E) YLDs. (F) standardized YLDs.

**Table 2.**
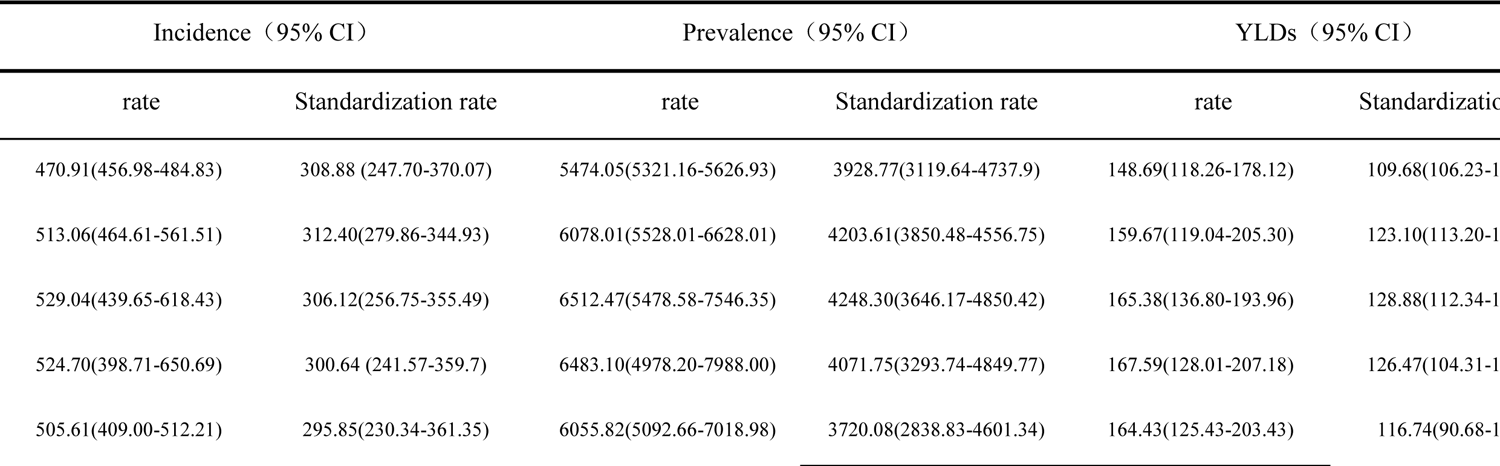
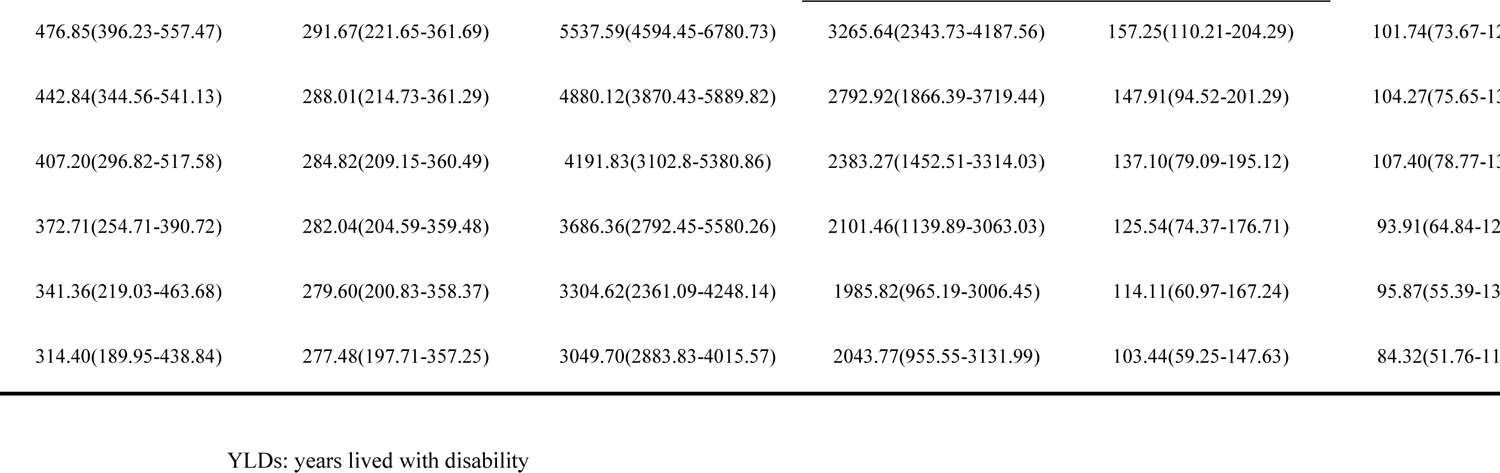
Projections of edentulism in China between 2020 and 2030 (1/100,000)

Finally, the standardized YLDs of missing teeth in China is projected to decrease from 90.52/100,000 to 84.32/100,000, a reduction of 6.85% when compared to 2019.

## 4 Discussion

Alongside economic development, there have been significant changes in the lifestyle of the population and a marked improvement in the standard of living over recent years; however, this has inevitably been accompanied by changes in the spectrum of human diseases. Over recent years, chronic and non-communicable diseases have become a key topic of global concern and increasing research attention has been paid to edentulism in different countries and regions. There has been a significant increase in research efforts in the basic, clinical, and public health fields related to edentulism, a phenomenon that rlects the significant health and economic burdens that chronic diseases impose on human society. As the most populous country, the prevention and treatment of edentulism in China has become a significant problem. Investigating the epidemiological distribution of edentulism and analyzing the trends of disease burden associated with edentulism over the next 10 years in large datasets generated domestically and internationally is very important if we are to develop methods to prevent and treat edentulism. Such studies can provide supportive data and the theoretical basis for the government to formulate prevention and control policies for edentulism.

In this study, we found that the disease burden associated with edentulism increased with age and that most of this burden was concentrated in individuals aged 70 years and over. The burden of disease was lowest in individuals aged 15–49 years, thus suggesting that changes in the age composition of the population exert changes in the trend of the disease burden for edentulism. The proportion of individuals in China aged 65 years and older increased from 26.32 million to 200 million between 1953 and 2021; this represents 4.4 to 14.2% of the population. This may be one of the main reasons for the overall increase in the disease burden associated with edentulism in China. This is because the risk of caries, periodontal disease, diabetes mellitus and other diseases associated with tooth loss increases with age, thus increasing the prevalence and incidence of tooth loss. Historically, the degree of ageing increased by an average of 0.15 and 0.18 per year from 1990 to 2000 and from 2000 to 2010, respectively, while the growth rate of population ageing accelerated significantly from 2010 to 2020, with an average annual growth rate of 0.46 [24]. This may explain the greater growth rate of the disease burden of edentulousness in China over recent years. In our study, the trend observed for age change was similar to that reported in our Fourth National Oral Health Survey[25], thus suggesting the validity and reliability of the study, which used GBD data. Although there are some differences in the disease burden of edentulism in terms of gender, previous studies have shown that because edentulism is associated with many factors, more studies are needed to confirm this despite the fact that the disease burden of edentulism is known to be higher in females than in males[14]; in the present study, we found that this difference was statistically significant [14].

In the present study, we used an ARIMA model for time series analysis to fit and predict the incidence, prevalence, and rate of YLDs in Chinese adults with rate and standardized rate edentulism. By performing these projections, we found that from 2020 to 2030, both rates and standardized rates are projected to first increase and then decrease, with an overall downwards trend. The incidence rate is projected to decrease from 470.91/100,000 to 314.40/100,000, the prevalence rate is projected to decrease from 5474.05/100,000 to 3049.70/100,000, and the YLDs is projected to decrease from 148.69/100,000 to 103.44/100,000. The standardized incidence is projected to decrease from 308.88/100,000 to 277.48/100,000. The standardized prevalence is expected to decrease from3928.77/100,000 to 2043.77/100,000 while the standardized YLDs is projected to decrease from 109.68/100,000 to 84.32/100,000. The relatively consistent downward trends in the rates and standardized rates suggest that the disease burden of edentulism in China will still follow a downwards trend by 2030, despite the further aging of the population.

The burden of edentulous disease is projected to rise between 2020 and 2023, possibly due to the fact that during the Coronavirus Disease (COVID-19) pandemic (2019-2023), dental services were not readily available to patients due to policy and personal factors; furthermore, patients did not generally attend hospitals for oral treatment except for emergencies such as traumatic injuries and acute endodontitis [26,32]. As a result, the incidence and prevalence of missing teeth are likely to increase as many teeth that could have been treated and preserved were not attended to in the clinic in a timely manner. In addition, the prevalence of caries and periodontal disease, the main causes of tooth loss, remain at a high level in China.

The disease burden associated with edentulism in China is projected to follow a decreasing trend between 2023 and 2030; this reduction is likely to be due to the introduction of a series of policies by the state. The National Health and Wellness Commission released the Healthy Dental Action Programme (2019–2025) on 31 January 2019, with separate age-appropriate programmes for the age-related high incidence of different diseases (caries, periodontal disease, and missing teeth).

In order to reduce the incidence of dental caries among adolescents, a special ‘sugar reduction’ campaign has been proposed; this requires primary and secondary schools, and childcare institutions, to restrict the sale of sugar-sweetened beverages and snacks, and to reduce the supply of sugar-sweetened beverages and sugar-sweetened foods in canteens [18,20]. Furthermore, the campaign promotes the implementation of intervention models for oral disease, such as children’s oral health check-ups, the prevention of caries by sealing grooves in the first molar, and the use of topical fluoride, by the health sector in conjunction with the education sector. There is a clear requirement to strengthen oral health and control the rate of caries in 12-year-old children to less than 25% by 2030. For middle-aged and elderly people, the prevention of periodontal disease is the main focus, and high-risk behavioral interventions for oral diseases are proposed. The construction of a smoke-free environment has been strengthened, the smoking ban in public places has been comprehensively promoted, and the supervision and enforcement of tobacco control in public places has been strictly enforced. In areas where the chewing of betel quid is a habit, efforts are focused on the oral health hazards of long-term betel quid chewing, targeted publicity and education, and oral health check-ups; this strategy will promote the early diagnosis and treatment of diseases such as periodontal and oral mucosal lesions. It is also important to promote the use of oral health-care products, such as health-care toothbrushes, fluoride-containing toothpaste and dental floss, promote the inclusion of oral health examinations in routine medical check-ups, and advocate the regular acceptance of oral health examinations, preventive oral cleaning, early treatments and other services for the prevention and treatment of oral diseases. It is also important to promote guidance and intervention for the prevention and treatment of oral diseases among the elderly, advocate that the elderly pay attention to the relationship between oral health and systemic health, strengthen the oral health management of patients with hypertension, diabetes and other chronic diseases among the elderly, and actively provide services such as the prevention and treatment of caries, periodontal diseases, oral mucosal diseases, and prosthetic denture repair. It is explicitly required that by 2025, the number of teeth retained by elderly people aged 65-74 years (in pieces) will reach 24.

President Xi Jinping proposed that we should ‘integrate health into all policies’ at the National Conference on Health and Health, which fully embodies our government’s policy of integrating oral health concepts into general health [34]. In response to the national policy, Shenzhen City has implemented production standards and set specifications for carbonated beverages from 1 January 2021, which explicitly requires that the shelves or counters of sugary beverages should be marked with added-sugar-related reminders [18,35]. Therefore, as a country showing high growth in sugar consumption, it is likely that China will also use consumption interventions to reduce sugar intake levels[36].

Moreover, forecasts indicate that China’s economic, scientific and technological strength, along with comprehensive national power, will attain a new levels between 2020 and 2030, and that the economy will embark on a road of development that is higher quality, more efficient, fairer, more sustainable, and more secure. Furthermore, it is expected that the economic growth rate will enter the ‘5 era’, with an average annual growth rate of approximately 5.3%. In addition, during the period of the Fourteenth Five-Year Plan (2021–2025), the average annual growth rate of China’s economy will reach approximately 5.3%; during the period of the Fifteenth Five-Year Plan (2026–2030), the growth rate will reach approximately 5.1% [37]. The relationship between socioeconomics and the disease burden of edentulism is complex [38]. On the one hand, many studies have found that a higher socioeconomic status is a protective factor against edentulism [39–41] and that the economy raises the level of education as well as cognitive ability. These factors are known to influence oral hygiene practices or the ways in which individuals interact with healthcare services, ultimately affecting the incidence and prevalence of edentulism. On the other hand, economic improvements at the national level may increase the types and quantities of ultra-processed foods and all types of beverages, thus increasing the prevalence of caries and periodontal disease, thereby also increasing the disease burden of edentulism[42]. In addition, some cultures, especially females of a higher socioeconomic status, may choose dentures because they are concerned about their appearance and the appearance of their teeth; this may, in part, increase the prevalence of edentulism in areas with better economic levels [38].

Although the initial stage of the introduction of the national policy has not been fully implemented, the burden of tooth loss disease in China between 2019 and 2023 is expected to follow an increasing trend. However, with the further implementation of the policy, and as the country’s future economic level continues to improve, the burden of tooth loss disease in China after 2023 is likely to follow a decreasing trend.

## Data Availability

This data was obtained from a publicly available web page

https://vizhub.healthdata.org/gbd-results/

