## Supplemental Table 1 for "Projecting trends in the disease burden of adult edentulism in China between 2020 and 2030: a systematic study based on the global burden of disease": Appendices.pdf

### Edentulism
### Flowchart

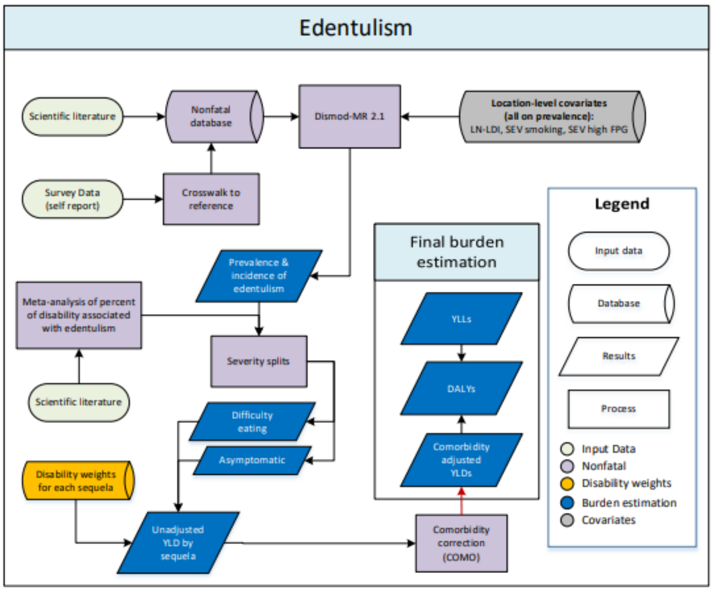


### Case definition

The case definition of edentulism includes any individual with zero remaining permanent teeth; toothlessness of infancy is not included. The assessment of this disease includes quantification of the prevalence of the disease as well as estimation of the major sequelae: asymptomatic toothlessness and symptomatic toothlessness leading to "great difficulty in eating meat, fruits, and vegetables." A small body of evidence has begun to emerge that implicates edentulousness as predisposing individuals to increased risk for ischaemic cardiovascular events including myocardial infarction and stroke. These data are sparse but have been included in models estimating the excess mortality of those with complete tooth loss. Given that the association is believed to be ecological rather than causal, however, edentulism has not been estimated as an underlying cause of death and it is not included in the risk factor analysis for cardiovascular diseases.

### Input data and data processing

Details of the systematic literature reviews are above. In addition to published studies, we also utilized self-report data on toothlessness from World Health Survey (WHS) for 47 countries as well as a number of national oral health surveys identified through the Global Health Data Exchange.

**Table 1: Total number of sources and countries with data for edentulism, by measure**

| | Total sources | Countries with data |
|---|---|---|
| All measures | 254 | 91 |
| Prevalence | 253 | 91 |
| Incidence | 1 | 1 |

### Age and sex splitting

The first step of data processing was age splitting. For any datum that did not entirely fit within a GBD sex or age group, the observation was split to be multiple age-specific and sex-specific data points based on the age and sex pattern predicted by previous DisMod-MR 2.1 models. It is our intention to update this age-sex splitting with each cycle of GBD.

### Crosswalks in MR-BRT

We then crosswalked self-reported (i.e. WHS) data on toothlessness to the reference definition of oral examination. In accordance with GBD 2019 principles for data processing, to make data comparable, we began by evaluating the number of observations of each alternate definition that matched with a corresponding observation from the reference definition. There were no “within” study matches identified so the MR-BRT analysis was based on 60 “between” study matches of alternative and reference definitions where a match was defined as data from different sources from the same GBD location, age group, and midpoint of the study period within 5 years of one another. The ratio of alternative to reference was calculated and logit-transformed. Standard error of the ratio was calculated using the delta method. Sex was included as a fixed effect and Socio-demographic Index (SDI) as a spline. The data matches, adjustment factors, and final input dataset are shown in the tables below.

**Table 2: Data points and matches between alternate and reference definitions**

| | Reference | Alternate #1 (cv_whs) |
|---|---|---|
| Number of data points | 10028 | 2874 |
| Within-study matches to reference | -- | 0 |
| Between-study matches to reference | -- | 60 |

**Table 3: MR-BRT Crosswalk Adjustment Factors for edentulism, 15% trim**

| Data input | Reference or alternative case definition | Gamma | Beta Coefficient, Logit (95% CI) | Adjustment factor* |
|---|---|---|---|---|
| Clinical exam | Reference | 0.019 | --- | --- |
| WHS (self-report) | Alt | | -0.126 (-0.416 to 0.154) | 0.882 (0.66 - 1.166) |

### Modelling strategy

Estimates for the prevalence of edentulism were calculated for each location/year/sex/age using DisMod-MR 2.1. As would be expected for an irreversible condition, remission was fixed at zero for all ages. Mortality and relative risk were both fixed at zero before age 30, as any excess cardiovascular events resulting from severe tooth loss would not be expected at younger ages. We also assigned incidence and prevalence to be zero during childhood. Incidence was allowed to rise beginning at age 15, which was chosen based on the age at which the permanent dentition is expected to have fully formed in all individuals. The random effect limits for all locations were bounded at +/- 1. As mentioned above, the criteria for diagnosis of edentulism are straightforward, and bias in the dataset was considered negligible. Thus, no study-level covariates were used in modelling the prevalence of edentulism. We included two location-level covariates in the model: 1) Log-transformed lag-distributed income (LDI) with a minimum beta value of 0.02 and 2) Log-transformed age-standardised summary exposure value (SEV) scalar of cardiovascular disease (CVD) in recognition of the common risk factors between CVD and tooth loss.

**Table 4: Covariate, parameter, beta, and exponentiated beta values for edentulism**

| Covariate | Param | Beta | Exponentiated beta |
|---|---|---|---|
| LN-LDI | Prev | -0.16 ( -0.17 — -0.16) | 0.85 (0.84 — 0.85) |
| SEV Smoking (age- and sex-specific) | Prev | 1.47 ( 1.34 — 1.60) | 4.36 (3.83 — 4.96) |
| SEV fasting plasma glucose (age- and sex-specific) | Prev | 0.25 ( 0.058 — 0.46) | 1.28 (1.06 — 1.58) |

Models were vetted based on the plausibility of the results, the extent to which estimates fit the data, and the plausibility of the range of estimates across location hierarchies.

### Severity distributions and disability weights

The disability weight used for symptomatic toothlessness leading to “great difficulty in eating meats, fruits, and vegetables” is 0.067 (0.045–0.095) as determined by the GBD disability survey. We considered all those with severe tooth loss and no access to dentures to experience this disability. However, the proportion of those with edentulism and severe tooth loss who have dentures has not been studied extensively. In order to estimate the proportion of edentulous individuals with no access to dentures, we completed a supplemental literature review of dentures prevalence for GBD 2010. Only six systematic surveys of dentures prevalence were identified, all in high- and middle-income countries. All were completed since 2000. After extracting the data from the studies, we performed linear regressions of denture presence and denture absence against health system access (HSA), a standardised covariate of treatment availability used in many disease estimation models. From the results of the regression, the prevalence of no dentures was calculated for all super-regions. We then completed a population-weighted average of all countries in the super-region based on 2003 populations, the average year of the dentures studies. Uncertainties for the prevalence of dentures were calculated by finding the standard deviation and standard error of the calculated prevalence values. The estimated prevalence of dentures in each location was used to calculate the proportion of individuals with asymptomatic edentulism and severe tooth loss (ie, those who have access to dentures) and difficulty eating due to edentulism and severe tooth loss (ie, those without access to dentures). This latter sequela was included as a cause of years lost due to disability (YLDs).

TableS1 Projections of edentulism in China, 2020-2030 (1/100,000)

| Incidence（95%CI） | | Prevalence（95%CI） | | YLDs（95%CI） | |
|---|---|---|---|---|---|
| Crude rate | Standardisation rate | Crude rate | Standardisation rate | Crude rate | Standardisatio[illegible] |
| 208.32 (160.05-261.99) | 284.56 (223.75-352.49) | 2324.02 (1794.69-3022.13) | 3520.03 (2737.47-4463.25) | 63.92 (40.76-96.90) | 95.59 (61.29-1[illegible] |
| 187.36 (145.30-235.34) | 257.00 (203.04-317.72) | 2057.87 (1601.32-2659.79) | 3101.10 (2426.08-3913.79) | 56.59 (36.17-85.70) | 84.15 (53.99-1[illegible] |

| | | | | | |
|---|---|---|---|---|---|
| 169.78 (132.58-213.92) | 233.67 (185.59-287.69) | 1826.65 (1422.55-2349.32) | 2740.78 (2146.68-3439.76) | 50.21 (31.97-75.92) | 74.31 (47.59-11[illegible] |
| 156.36 (121.87-197.80) | 215.38 (170.59-266.54) | 1644.85 (1284.62-2107.51) | 2456.40 (1918.69-3086.06) | 45.19 (28.74-68.28) | 66.54 (42.52-9[illegible] |
| 148.06 (114.75-188.03) | 202.95 (160.78-251.30) | 1529.53 (1195.66-1955.14) | 2265.09 (1777.60-2846.93) | 41.99 (26.58-63.29) | 61.30 (39.05- 9[illegible] |
| 145.64 (112.43-185.10) | 197.22 (155.42-244.93) | 1497.79 (1171.79-1910.97) | 2184.35 (1718.06-2773.17) | 41.10 (25.99-61.99) | 59.09 (37.77-8[illegible] |
| 150.41(115.54-191.14) | 198.93 (156.70-247.98) | 1552.07 (1210.90-1976.45) | 2208.82 (1725.49-2807.51) | 42.58 (27.03-63.90) | 59.77 (38.17-8[illegible] |
| 160.75 (122.39-205.33) | 206.18 (160.51- 257.62) | 1663.34 (1289.14-2123.86) | 2298.73 (1777.06-2929.99) | 45.62 (28.83-68.58) | 62.23 (39.55-9[illegible] |
| 173.56 (131.76-222.03) | 215.81 (165.31- 270.97) | 1805.16 (1387.65-2309.83) | 2419.32 (1861.93-3098.77) | 49.50 (31.39-74.50) | 65.52 (41.72- 9[illegible] |
| 185.76 (139.75-238.20) | 224.54 (171.68-282.50) | 1948.64 (1485.89-2497.03) | 2534.76 (1938.93-3251.04) | 53.43 (33.85-80.69) | 68.68 (43.73-10[illegible] |
| 193.76 (145.37-246.64) | 229.08 (174.63-289.26) | 2058.56 (1560.32-2638.05) | 2608.85 (1986.52-3342.15) | 56.42 (35.65-85.45) | 70.71 (44.77-10[illegible] |
| 201.42 (151.22-256.40) | 232.61 (177.68-292.94) | 2167.27 (1645.94-2776.04) | 2678.02 (2047.46-3421.20) | 59.41 (37.85-90.26) | 72.61 (46.20-10[illegible] |
| 212.90 (161.51-269.80) | 239.22 (183.73-300.15) | 2316.07 (1778.71-2964.15) | 2786.26 (2140.68-3521.02) | 63.50 (40.54-96.34) | 75.58 (47.88-11[illegible] |
| 226.22 (173.57-285.60) | 246.84 (191.90-307.94) | 2485.29 (1921.44-3174.97) | 2908.24 (2256.23-3663.72) | 68.18 (43.40-103.27) | 78.95 (50.33-11[illegible] |
| 239.49 (185.40-301.30) | 253.40 (199.63-315.45) | 2655.52 (2060.76-3396.80) | 3018.47 (2343.69-3807.97) | 72.88 (46.40-109.88) | 81.98 (52.30-12[illegible] |
| 249.48 (194.72-313.28) | 256.89 (202.64-319.02) | 2795.49 (2160.71-3583.00) | 3092.05 (2411.49-3900.85) | 76.73 (49.05-115.29) | 84.01 (53.43-12[illegible] |
| 259.54 (202.75-326.15) | 259.83 (205.04- 322.65) | 2940.44 (2286.78-3760.54) | 3156.76 (2456.88-3981.99) | 80.74 (51.54-121.43) | 85.83 (54.56-12[illegible] |
| 272.46(212.75-342.64) | 264.96 (207.90-329.24) | 3117.85 (2430.29-3981.75) | 3243.69 (2522.50-4092.00) | 85.65 (55.00-128.76) | 88.27 (56.19-13[illegible] |
| 286.26 (223.31-360.29) | 270.54 (212.18-336.46) | 3303.04 (2564.43-4215.04) | 3332.13 (2588.27-4204.04) | 90.78 (58.33-136.47) | 90.77 (57.97-13[illegible] |
| 299.40 (232.87-376.94) | 274.76 (214.95-342.10) | 3475.43 (2685.30-4434.83) | 3400.28 (2642.77-4296.05) | 95.56 (61.23-143.41) | 92.70 (59.05-13[illegible] |
| 308.76 (239.60-388.24) | 275.91 (215.46-343.27) | 3600.18 (2778.49-4598.27) | 3427.00 (2658.96-4333.29) | 99.00 (63.00-148.75) | 93.47 (59.57-13[illegible] |
| 306.47 (237.12-387.54) | 267.07 (208.88-333.20) | 3575.03 (2768.34-4551.35) | 3311.61 (2575.52-4180.95) | 98.29 (62.78-147.59) | 90.31 (57.63-13[illegible] |
| 289.34 (223.96-364.77) | 247.43 (194.29-307.73) | 3360.95 (2618.63-4256.72) | 3037.76 (2376.28-3809.25) | 92.37 (59.33-138.95) | 82.80 (53.00-12[illegible] |
| 265.89 (207.03-334.35) | 224.21 (178.07-277.53) | 3065.47 (2398.30-3865.47) | 2709.49 (2121.40-3369.28) | 84.22 (53.65-126.35) | 73.81 (47.24-10[illegible] |
| 246.07 (191.66-309.65) | 204.59 (162.69-254.87) | 2813.40 (2199.19-3540.36) | 2430.14 (1908.46-3037.53) | 77.25 (48.81-115.79) | 66.16 (42.24-9[illegible] |
| 239.89 (186.50-303.10) | 195.82 (155.40-244.28) | 2732.10 (2140.98-3431.34) | 2304.18 (1810.75-2872.02) | 74.98 (47.34-112.29) | 62.71 (40.00-9[illegible] |
| 245.09 (190.44-309.92) | 194.92 (154.26-242.85) | 2792.50 (2187.15-3510.49) | 2292.11 (1806.18-2854.58) | 76.58 (48.70-114.40) | 62.36 (39.80-9[illegible] |
| 251.20 (194.43-318.02) | 194.58 (154.01-242.16) | 2863.93 (2240.96-3609.08) | 2288.72(1802.00-2851.63) | 78.48 (50.05-117.33) | 62.26 (39.77-9[illegible] |
| 295.41 (229.39-371.58) | 218.80 (173.76-269.75) | 3390.30 (2653.02-4313.06) | 2613.10 (2047.35-3259.17) | 92.82 (59.43-140.27) | 71.08 (45.58-10[illegible] |

| | | | | | |
|---|---|---|---|---|---|
| 388.93 (298.29-491.58) | 272.15 (214.34-337.96) | 4512.78 (3485.26-5830.61) | 3327.48 (2589.33-4226.85) | 123.44 (79.31-186.25) | 90.52 (57.75-13[illegible] |
